# Perinatal risk factors, DNA methylation and the development of ADHD symptoms: a high-dimensional mediation analysis

**DOI:** 10.64898/2026.08.10.26360078

**Authors:** Alexander Neumann, Matthew Suderman, Janine F Felix, Charlotte Cecil

## Abstract

**Background:** Attention-deficit/hyperactivity disorder (ADHD) is associated with perinatal and genetic risk factors, including prenatal maternal smoking, pre-pregnancy BMI, gestational age, birth weight, and common genetic variants. These risk factors, as well as ADHD symptoms themselves, have previously been linked to cord blood DNA methylation (DNAm). We tested the hypothesis that cord blood DNAm mediates the effects of these risk factors on ADHD symptoms.

**Methods:** Participants were drawn from two large European population-based cohorts: the Generation R Study and Avon Longitudinal Study of Parents and Children (n=3087). Cord blood DNAm was assessed using Illumina 450k and EPIC v1 arrays. ADHD symptoms were repeatedly measured with parent-based questionnaires between the ages 6 and 10 years. A high-dimensional mediational model based on DNAm principal components mediation analysis (PCMA) estimated the global mediation effect of all tested DNAm sites. Mediation via single principal components and individual DNAm sites was also evaluated using structural equation modeling and Divide-Aggregate Composite-null Test (DACT).

**Results:** DNAm globally mediated the relationships of maternal smoking, low birth weight, and an ADHD polygenic score (PGS) with ADHD symptoms. Specifically, DNAm explained 62% of the total effect for maternal smoking, 56% for birth weight, and 35% for the ADHD-PGS. No association with individual principal components or single DNAm sites survived multiple testing correction. Evidence for mediation was absent for pre-pregnancy BMI and inconsistent for gestational age.

**Conclusions:** In this first epigenome-wide mediation study of ADHD, we demonstrate a role of DNAm at birth in mediating the association of maternal smoking, birth weight and ADHD-related genetic variants with ADHD symptoms. However, lack of individual site-specific findings and the observational design limit causal biological interpretations. We therefore encourage further research of epigenetic pathways for these three risk factors.

## Introduction

Attention-deficit and hyperactivity disorder (ADHD) is a neurodevelopmental disorder characterized by attention problems, impulsivity and excessive activity. The disorder has an early peak age-of-onset of 9.5 years (Solmi et al., 2022), affecting approximately 5-7.5% of children worldwide (Faraone et al., 2021; Thomas et al., 2015). Etiological ADHD research points to a role of both genetics and the early environment.

Molecular (van der Laan et al., 2025) and family genetic (Larsson et al., 2014; Taylor et al., 2023) studies suggest a substantial genetic component, with twin heritability estimates between 66-88%. Beyond genetics, numerous environmental risk factors have been linked to ADHD development. Perinatal risk factors are of particular interest due to the early development of the disorder, the importance of this period to neural development and the potential for early intervention. Among others, maternal smoking (Kim et al., 2020; Tsegay et al., 2026; Zakerinasab et al., 2026), higher pre-pregnancy BMI (Kim et al., 2020; Li et al., 2020; Park et al., 2026), lower gestational age (Ask et al., 2018; Nivins et al., 2026; Salontaji et al., 2024) and lower birth weight (Heinonen et al., 2010; Momany et al., 2018) have consistently been associated with ADHD. While each risk factor may operate through distinct biological mechanisms, epigenetic modification is a hypothesized shared pathway (Momany et al., 2018; Park et al., 2026; Salontaji et al., 2024; Tsegay et al., 2026). All four perinatal risk factors strongly associate with DNA methylation (DNAm) in cord blood, with a hundred or more DNAm sites identified at birth in epigenome-wide meta-analytic studies (EWASs) for each (Joubert et al., 2016; Küpers et al., 2019; Merid et al., 2020; Sharp et al., 2017). Interestingly, genetic predisposition for ADHD appears to also be related to epigenetic profiles at birth: examining cord blood DNAm and a polygenic score (PGS) for ADHD, we recently reported that 166 differentially methylated regions were associated with an ADHD-PGS (Schuurmans et al., 2025).

DNAm at birth is not only linked to perinatal exposures, but also to ADHD symptoms in childhood. Several DNAm sites in cord blood (Neumann et al., 2020; Walton et al., 2017) or neonatal buccal samples (Camerota et al., 2024) have been associated with ADHD symptoms. We previously observed substantial specificity to the birth period: DNAm effects at birth only modestly correlated with those identified cross-sectionally in childhood (Neumann et al., 2025). We thus hypothesize that cord blood DNAm in particular may represent perinatal exposures relevant for ADHD. The observation that both perinatal exposures and ADHD are related to DNAm at birth naturally raises the question whether DNAm has a role in mediating the risk effects of perinatal exposures on ADHD. Cord blood DNAm levels altered by perinatal exposures could directly influence ADHD risk, e.g. via systemic effects like inflammation, or could serve as a proxy for another causal mechanisms, e.g. brain methylation directly involved in neural development. Since observational data cannot readily distinguish these different mechanisms, we use “mediation effects” to describe all forms of statistical mediation.

In summary, epigenetic mechanisms are an often proposed mechanism to explain the risk effects of perinatal factors, but formal tests of mediation have hardly been performed. A potential reason for this research gap might be the low power of mediation tests due to a composite null, i.e. a null effect which can arise due to both a non-significant a or b path (Barfield et al., 2017). To circumvent this, researchers can estimate the joint mediation effects of multiple DNAm sites simultaneously using high-dimensional mediation models (Clark-Boucher et al., 2023). Alternatively, post-hoc p-value adjustments can be applied using methods like the Divide-Aggregate Composite-null Test (DACT) (Liu et al., 2022; Zeng et al., 2021).

In this study our goal was I) to quantify and test the global mediation effect, i.e. how much, in total, does DNAm across the genome mediate the risk effects of perinatal and genetic risk factors on ADHD symptoms; and II) to characterize the most important DNAm sites underlying potential global mediation effects. Due to the high-dimensionality of DNAm relative to sample size, we will evaluate mediation using Principal Component Mediation Analysis (PCMA; Huang & Pan, 2016) and apply it to repeated measures of ADHD in two large European population-based cohorts (n=3087): the Generation R Study (GenR) and the Avon Longitudinal Study of Parents and Children (ALSPAC).

## Methods

### Participants

An overview of the participant selection, data preparation and statistical methods can be found in Figure 1.

**Figure 1:**
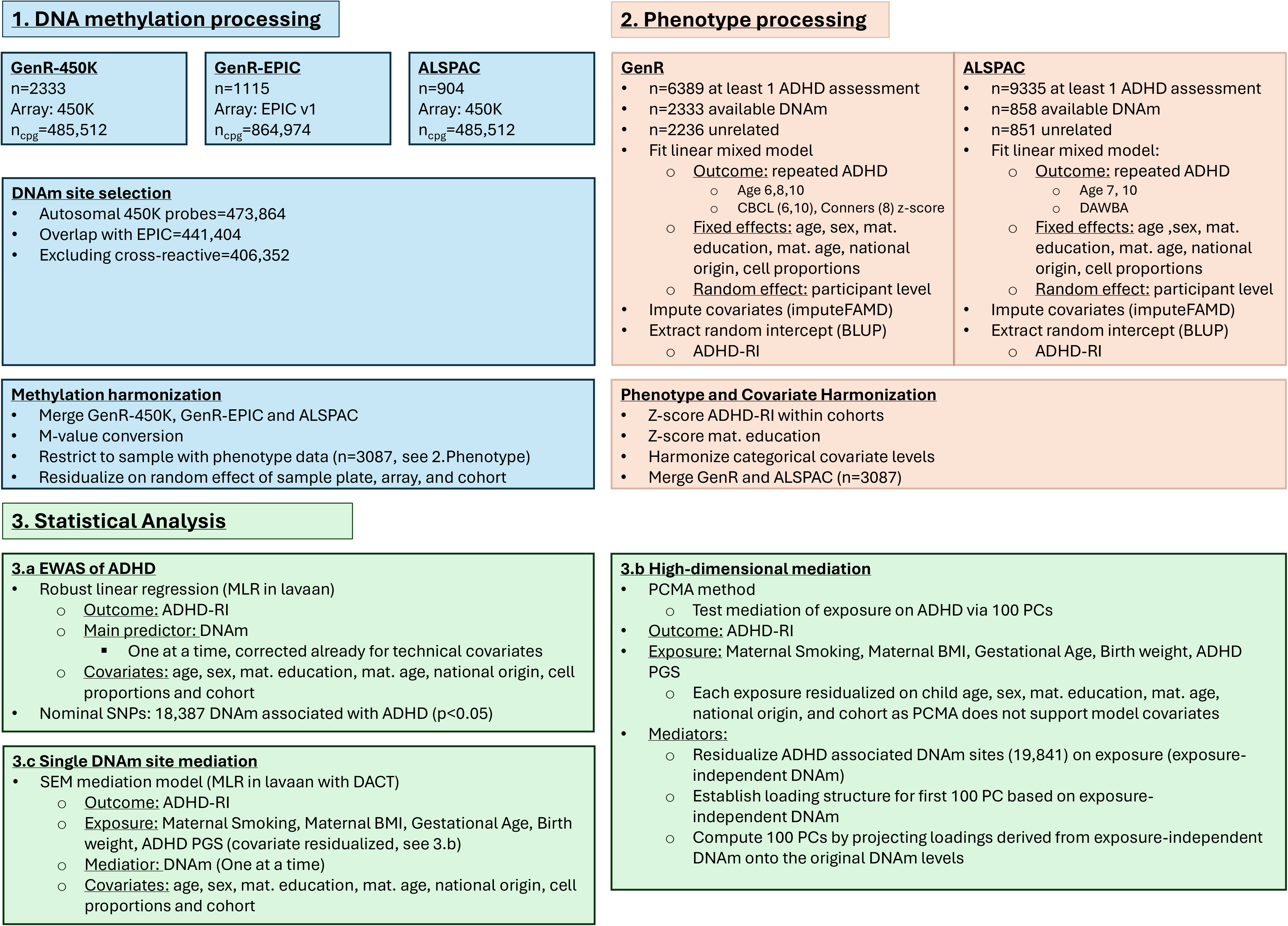
Methods overview.

#### Generation R Study

GenR is an ongoing population-based birth cohort following parents and children from fetal life. Between April 2002 and January 2006, 9,778 pregnant residents of Rotterdam, the Netherlands, were enrolled. For 6389 participants ADHD was assessed at least once at either age 6, 8 or 10 years. Out of these, DNAm at birth was available for 2333 children. In case of related participants one sibling was excluded based on data availability or, if equal, randomly, to ensure independent observations. The resulting analysis sample was 2236 participants.

#### ALSPAC

Between April 1991 and December 1992 eligible pregnant women in Avon, UK (20,248 pregnancies) were invited to take part in ALSPAC. Initial number of pregnancies enrolled was 14,541. Out of 14,676 fetuses, 14,062 were live births and 13,988 infants survived to age one. See previous publications for a full cohort description (Boyd et al., 2013; Fraser et al., 2012). 9335 participants had at least one ADHD assessment at either age 7 or 10, and of these 858 had DNAm measured at birth. Seven siblings were excluded resulting in final analysis sample of 851 participants. Please note that the study website contains details of all the data that is available through a fully searchable data dictionary and variable search tool (http://www.bristol.ac.uk/alspac/researchers/our-data/).

#### Ethical approval

GenR was approved by the Erasmus MC Medical Ethical Committee (MEC 198.782.2001.31) and ALSPAC by the ALSPAC Ethics and Law Committee and the Local Research Ethics Committees. All participating families provided written informed consent, see supporting information.

### Measures

#### Perinatal risk factors

Maternal smoking during pregnancy was assessed with questionnaires (Duijts et al., 2008; Richmond et al., 2014). We categorized smoking as any versus no smoking during pregnancy, see supporting information and Table S1 for comparisons with sustained/stopping smoking. Pre-pregnancy BMI was calculated from measured height at intake and self-reported weight before pregnancy. In GenR, missing pre-pregnancy BMI was regression imputed using BMI in the first trimester. Gestational age was based on ultrasound in GenR (Verburg et al., 2008) and in ALSPAC mostly on last menstrual period (see supplemental information). Birth weight was obtained from birth records.

#### ADHD-PGS

We used an existing PGS from previous work (Schuurmans et al., 2025) based on an ADHD case-control GWAS (Demontis et al., 2023) and computed using a clumping and thresholding approach with PRSice-2 (Choi & O’Reilly, 2019). The PGS was z-scored and residualized for twenty genomic principal components to account for population stratification. See supporting information for genotyping details.

#### DNA methylation

DNAm was measured in cord blood with either the Illumina HumanMethylation450 BeadChip array in GenR (GenR-450K) and ALSPAC (ALSPAC-450K) or MethylationEPIC v1.0 BeadChip in GenR only (GenR-EPIC). See supporting information and previous publications for details on pre-processing (Mulder et al., 2021). We restricted the DNAm matrix to autosomal sites present on both arrays. We excluded cross-reactive probes using the maxprobes package(Chen, 2018/2024). Methylation values were converted to M-values. We merged the GenR-450K, GenR-EPIC and ALSPAC-450K datasets and residualized M-values on random effects of sample plate, array and cohort to reduce technical and population-specific effect, as PCMA does not support covariate inclusion in the analysis model.

#### ADHD measures

In GenR, ADHD symptoms were measured using parent-rated questionnaires. Specifically, at age 6 years with the Child Behavior Checklist (CBLC) for ages 1.5-5 (Achenbach & Rescorla, 2000; Achenbach & Ruffle, 2000), at age 8 years with the Conners’ Parent Rating Scale (CPRS-R)(Conners et al., 1998), and at age 10 years with the CBLC for ages 6-18 (Achenbach & Rescorla, 2001). To harmonize the different questionnaires, each ADHD subscale was z-score standardized. In ALSPAC, the Development and Well-Being Assessment (DAWBA)(Goodman et al., 2011) was administered to parents at age 7 and 11 years. This instrument generates probability bands ranging from 0 to 5, indicating very low probability of ADHD (<0.1% of children) to very high probability (>70% of children). The choice of timepoints and measures was informed by a previous multi-cohort EWAS including GenR and ALSPAC, which successfully identified DNAm sites at birth associated with these repeated ADHD outcomes (Neumann et al., 2020).

#### Covariates

DNAm was residualized for sample plate, array and cohort (see DNA methylation). In addition, all mediation pathways are adjusted for age at ADHD assessment, child sex (genetically confirmed), maternal education, maternal age at intake, national origin (European yes/no), and cell proportions. Maternal education was defined as continuous variable ranging from no education to higher education and z-score standardized within GenR and ALSPAC. Cell proportions were estimated using the Houseman method with a combined reference panel for cord blood (Gervin et al., 2019). Granulocytes were excluded/set as reference to avoid multi-collinearity. In secondary analyses, to account for genetic confounding, we additionally adjusted for the ADHD-PGS.

### Statistical Analysis

We tested the hypothesis that perinatal and genetic risk effects on ADHD are mediated via DNAm using PCMA, a high-dimensional mediation model able to estimate the joint mediation effect of DNAm sites across the genome (Huang & Pan, 2016). All analyses were conducted with R 4.5.1 (R Core Team, 2022). Several preparatory steps were conducted before PCMA: 1) account for repeated measures, 2) impute missing data, 3) estimate the total effect and 4) screen DNAm sites (Figure 1).

In this study we extended a longitudinal random effects model used in a previous ADHD EWAS (Neumann et al., 2020). However, as PCMA does not support random effects, we applied a two-step approach. We first fitted a linear mixed model with lme4 (Bates et al., 2014), regressing repeated measures of ADHD within either GenR or ALSPAC on covariates (fixed effects) and the participant level as random intercept. We extracted the best linear unbiased predictor of the random intercept, which represents participant-specific ADHD levels across childhood conditional on the covariates (ADHD-RI). Additionally, we harmonized ADHD-RI between cohorts by calculating z-scores within each cohort. Missing outcome data was accounted for with ML.

Next, to reduce selection bias, maximize power and allow comparability between exposures, we imputed missing predictors and covariate data (missing proportion < 4%, Table 1). We used imputeFAMD in missMDA 1.20, a PCA-based implementation of single imputation (Josse & Husson, 2016). Number of PCs was determined using 10-fold cross-validation. After imputation, GenR and ALSPAC data were merged for mega-analysis.

**Table 1:**
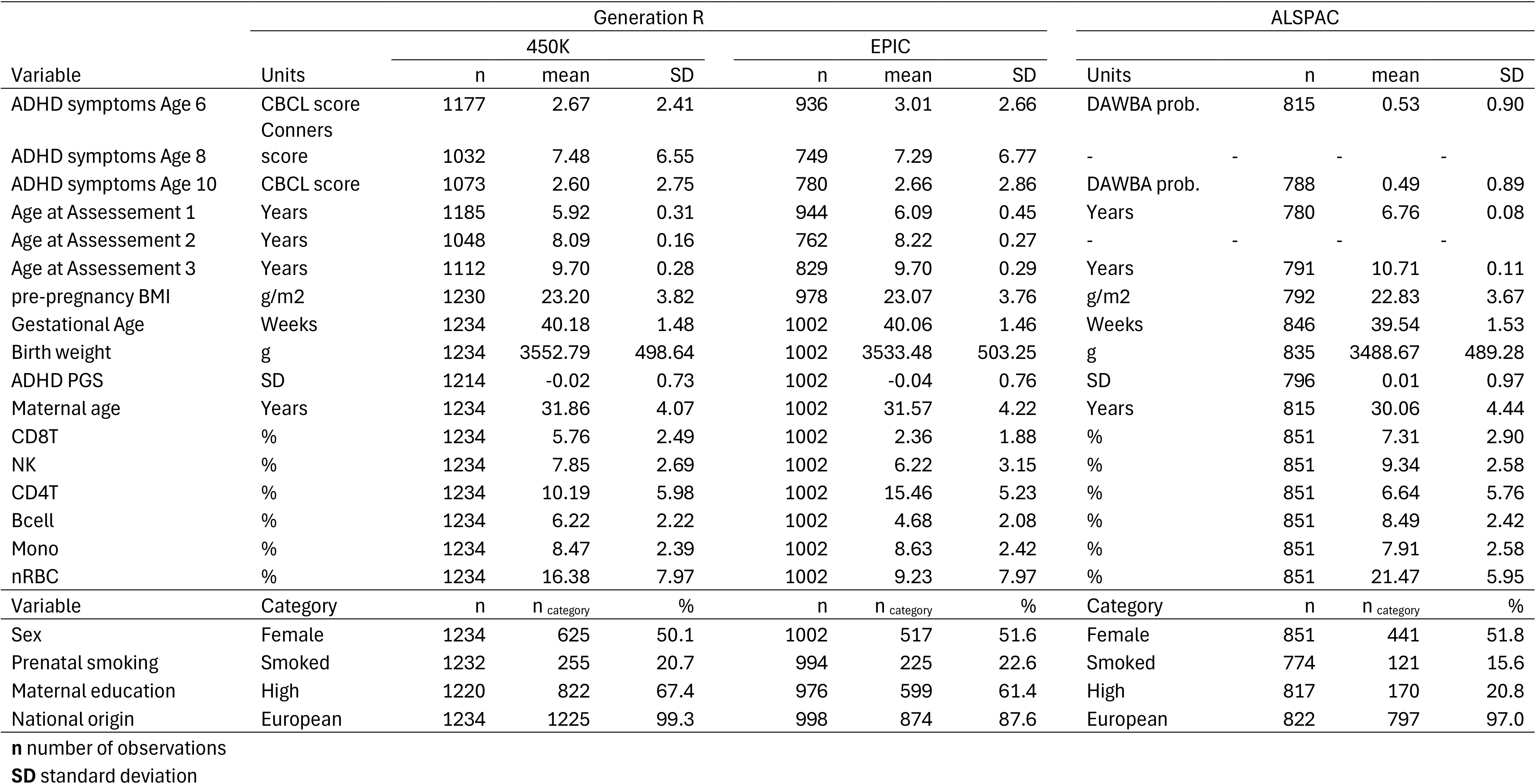
Descriptives.

Before running the mediation models, we first established whether the exposures associate with ADHD (total effect). We regressed ADHD-RI on perinatal risk factors in separate models, as well as in a joint model to explore independent associations. The predictors were residualized on covariates, for consistency with PCMA.

Another preparatory step included pre-selecting DNAm sites which associate with ADHD at nominal significance. High-dimensional mediation models aim to estimate genome-wide effects, yet they cannot handle more than several hundred mediators at one time due to statistical and computational limitations (Clark-Boucher et al., 2023). We screened for association with ADHD symptoms to ensure the same selection of DNAm sites for each exposure. We regressed ADHD-RI on each DNAm site adjusting for covariates using robust ML in Lavaan 0.6-21 (Rosseel, 2012). We then selected nominally significant DNAm sites (p<0.05) for further consideration.

After screening, to allow comparisons exposure variables across both cohorts were z-scored, except for the ADHD-PGS which was already standardized within genotype dataset. We also residualized the risk factors on the covariates, as PCMA does not support covariate adjustment in the model. As DNAm and ADHD-RI are already conditional on covariates, all paths are controlled for potential confounders.

Finally, mediation was tested with PCMA, using the mcma_PCA function in spcma 1.0 (Zhao et al., 2020). PCMA relies on testing mediation via DNAm PCs (Huang & Pan, 2016). Specifically, the loading structure of the PCs is established based on DNAm levels residualized for the exposure to avoid overfitting the PCs in a way that they merely become a biomarker of the exposure. The loading structure is then projected onto the original DNAm levels. We applied PCMA to the top 100 PCs, previously shown to capture global meditation effects in simulations well (Clark-Boucher et al., 2023). Next the mediation effects of all 100 PCs, by definition uncorrelated, are estimated simultaneously. This enables the estimation of mediation effects via individual PCs and the summation of these effects allows the estimation of a global DNAm effect, which are tested using quasi-Bayesian approximation. Individual PC results were corrected for false discovery rate (FDR).

PCs showing at least nominally significant mediation were further characterized by testing whether the top 1000 loading DNAm sites were enriched for GO biological and KEGG pathways. Enrichment was tested with gometh in missMethyl 1.44 (Maksimovic et al., 2021). We selected only nominally significant GO pathways and removed redundant terms with reducesimmatrix from rrvgo 1.22 (Sayols, 2023).

To complement PC-based mediation, we also applied traditional single DNAm site mediation models in Lavaan with the same covariates and MLR. The p-value for the mediation effect was computed using the DACT method, which prevents deflation of mediation test statistics in genome-wide studies.(Liu et al., 2022) Analysis code can be found at github.com/aneumann-science/perinatal_mediation.

## Results

GenR and ALSPAC showed similar levels of exposures and covariates, except for maternal smoking and maternal education, which were both higher in GenR (Table 1). The risk factors showed positive, but mostly low intercorrelations (r<0.12), except for gestational age and birth weight (r=0.46) (Table 2). The ADHD-PGS correlated r=0.05 with maternal smoking and below r=0.03 with the other factors.

**Table 2:** Correlations betweens perinatal risk factors adjusted for covariates.

|  | Maternal smoking | Pre-pregnancy BMI | Lower gestational age | Lower birth weight | ADHD PGS |
| --- | --- | --- | --- | --- | --- |
| Maternal smoking | 1.00 | -0.03 | 0.00 | 0.06 | 0.05 |
| Pre-pregnancy BMI | -0.03 | 1.00 | -0.01 | -0.12 | 0.02 |
| Lower gestational age | 0.00 | -0.01 | 1.00 | 0.46 | 0.01 |
| Lower birth weight | 0.06 | -0.12 | 0.46 | 1.00 | 0.00 |
| ADHD PGS | 0.05 | 0.02 | 0.01 | 0.00 | 1.00 |

Pearson correlations between perinatal risk factors. All factors are residualized for child age at assessment, sex, maternal education, maternal age, national origin, cell proportions

We next estimated the total effect of the perinatal risk factors on ADHD. All four perinatal exposures showed significant associations with similar standardized coefficients ranging from 0.04SD (95% CI: [0.00; 0.08], p=0.03) for gestational age to 0.06SD for maternal smoking ([0.03; 0.10], p=7.3*10^-4^) (Table 3). These associations were largely independent of each other, except for gestational age – a mutually adjusted model suggested that almost half of the explained variance can be ascribed to birth weight. The ADHD-PGS showed higher total effects on ADHD symptoms compared to the environmental factors: A 1-SD higher PGS was associated with 0.17SD higher ADHD scores ([0.13; 0.22], p=6.9*10^-14^). The total effects of the perinatal risk factors was largely independent of the ADHD-PGS.

**Table 3:**
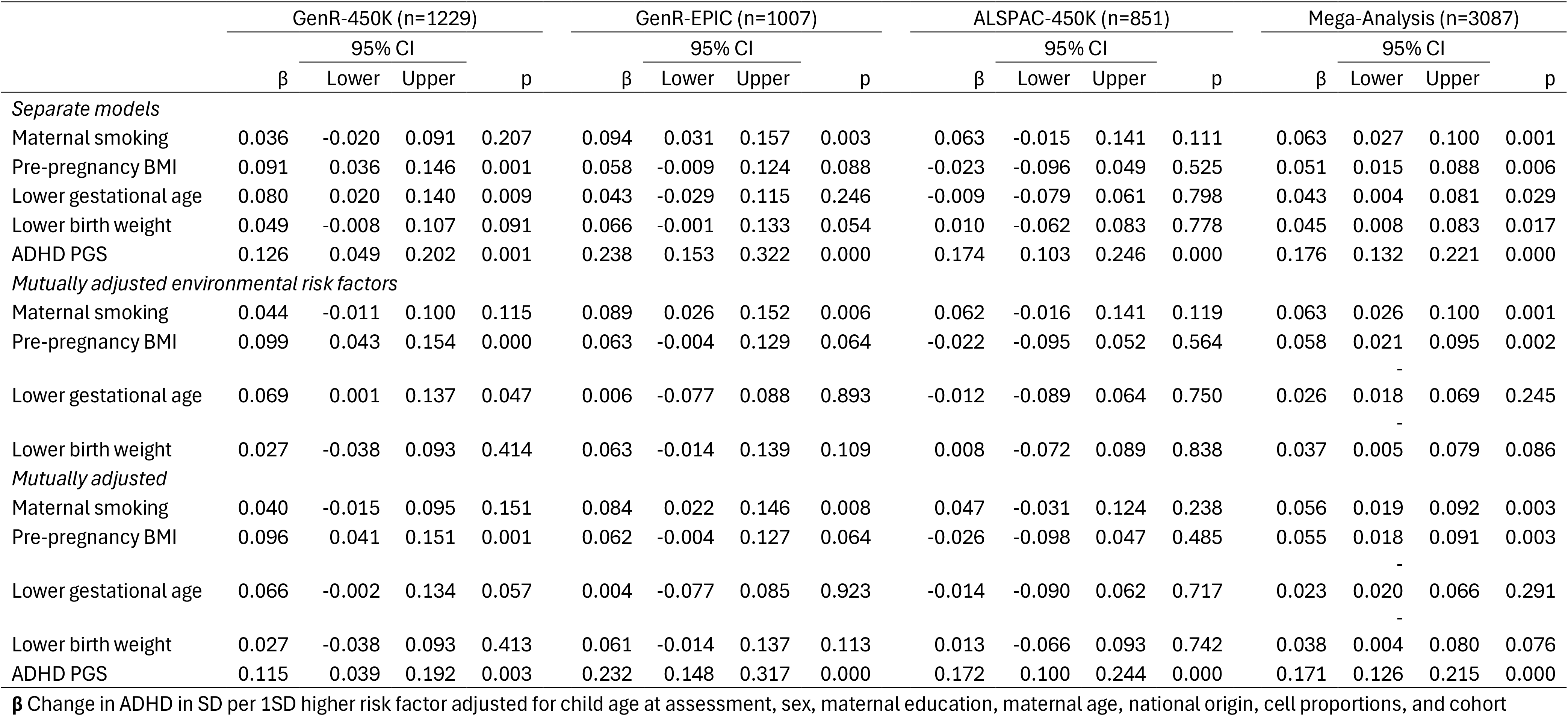
Total effect of perinatal risk factors.

Before continuing with the mediation models, we screened DNAm sites for nominal (p<0.05) associations with ADHD. We identified 18,387 DNAm sites nominally associated and considered them further in high-dimensional mediation analyses.

Next, we estimated the global meditation effect, i.e. the joint mediation effect of all 100 methylation PCs based on the 18,387 ADHD-associated DNAm sites (Figure 2, Table S2). DNAm jointly mediated 61.9% of the overall effect of maternal smoking on ADHD symptoms (β_indirect_=0.039, [0.016, 0.062], p<0.001) with consistent mediation effects estimated in all (sub-)cohorts. The top PC contributing to this mediation effect was PC3 with a proportion mediated of 5.7%, but it did not pass multiple testing correction (p=0.02, q=0.98) (Table 4). Pathway enrichment analyses of the top 1000 loading DNAm sites did not survive adjustment for multiple testing for any of the examined PCs, however, top pathways included cornified envelope formation (p=1.4*10^-4^, q=0.05) and sebaceous gland development (p=7.2*10^-5^, q=1.00) (Table 5).

**Figure 2:**
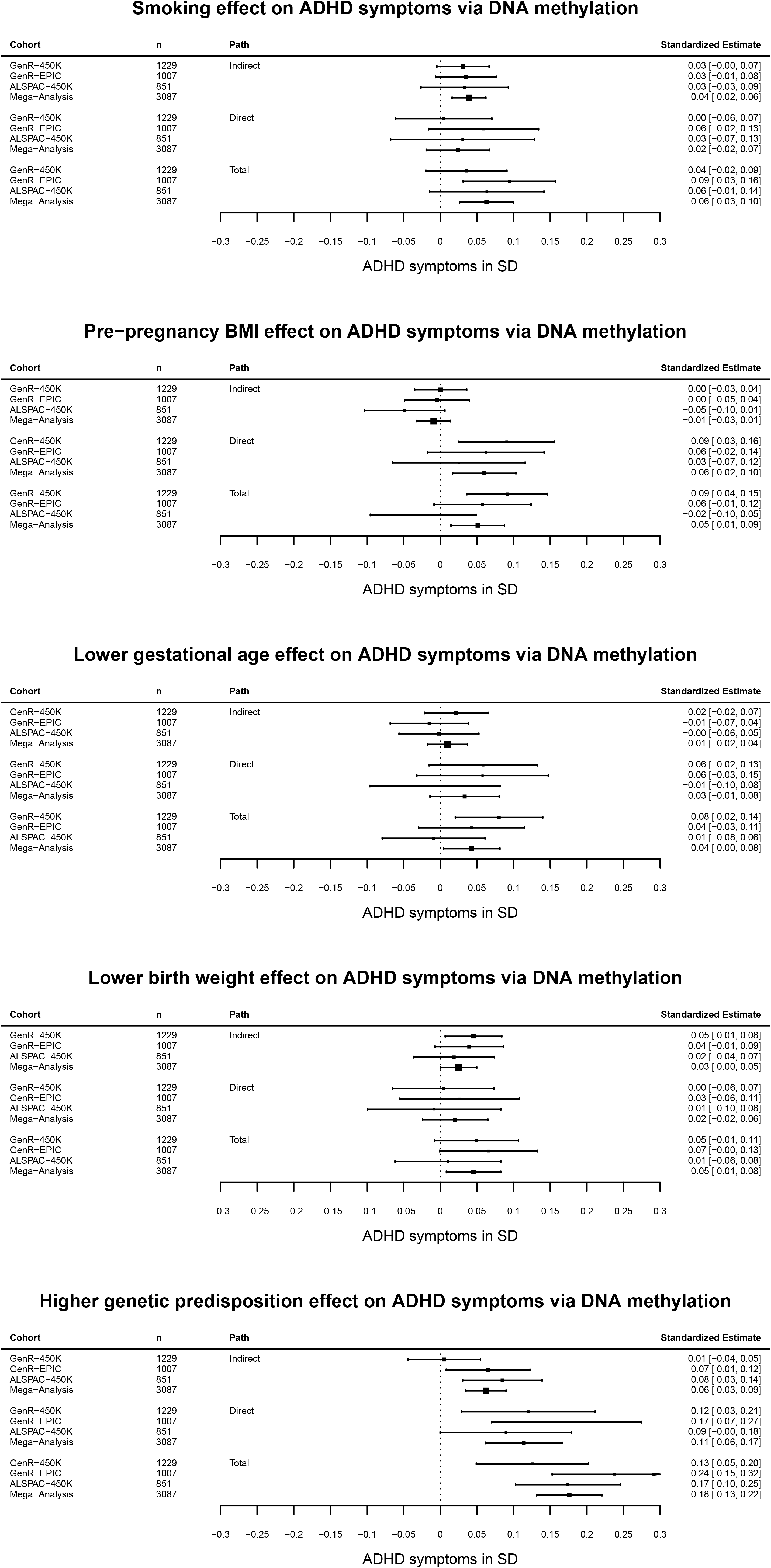
Global mediation estimates of perinatal risk factor effects on ADHD symptoms via DNAm. Global mediation estimates are based on PCMA analyses. They represent the standardized effect of a perinatal risk factor via 100 DNAm PCs, which themselves are based on 18,387 DNA methylation sites nominally associated with ADHD symptoms. **“**Indirect” path refers to the mediated effect (a*b), “Direct” to the non-mediated pathway (c’) and “Total” to the effect of both pathways (a*b+c’). Estimates are provided across the whole study sample (Mega-analysis), as well as stratified by (sub-)cohort. Analyses were adjusted for child age, sex, maternal education, maternal age, national origin, and cohort. DNAm was residualized for sample plate, array, and cohort.

**Table 4:**
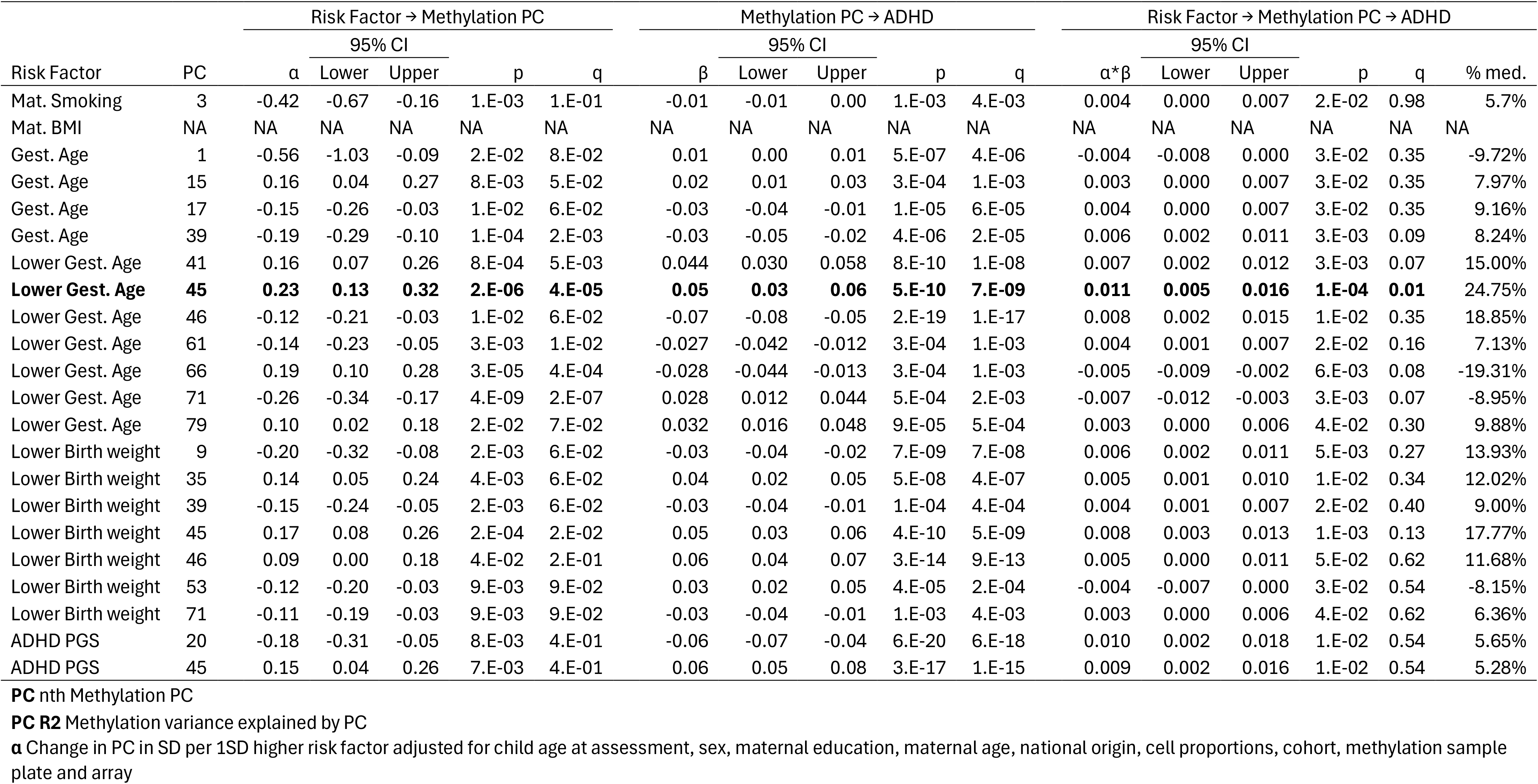
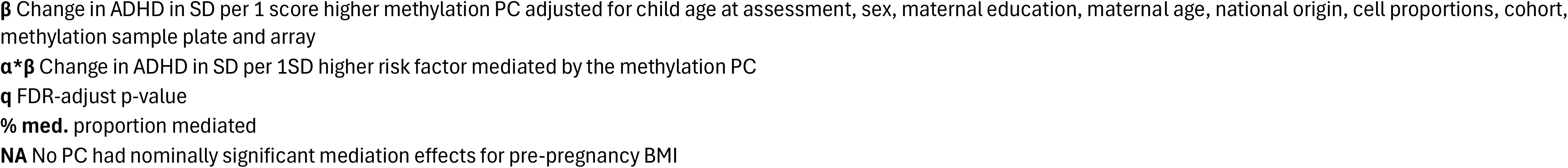
Mediation individual PCs with nominally significant mediation effects.

**Table 5.**
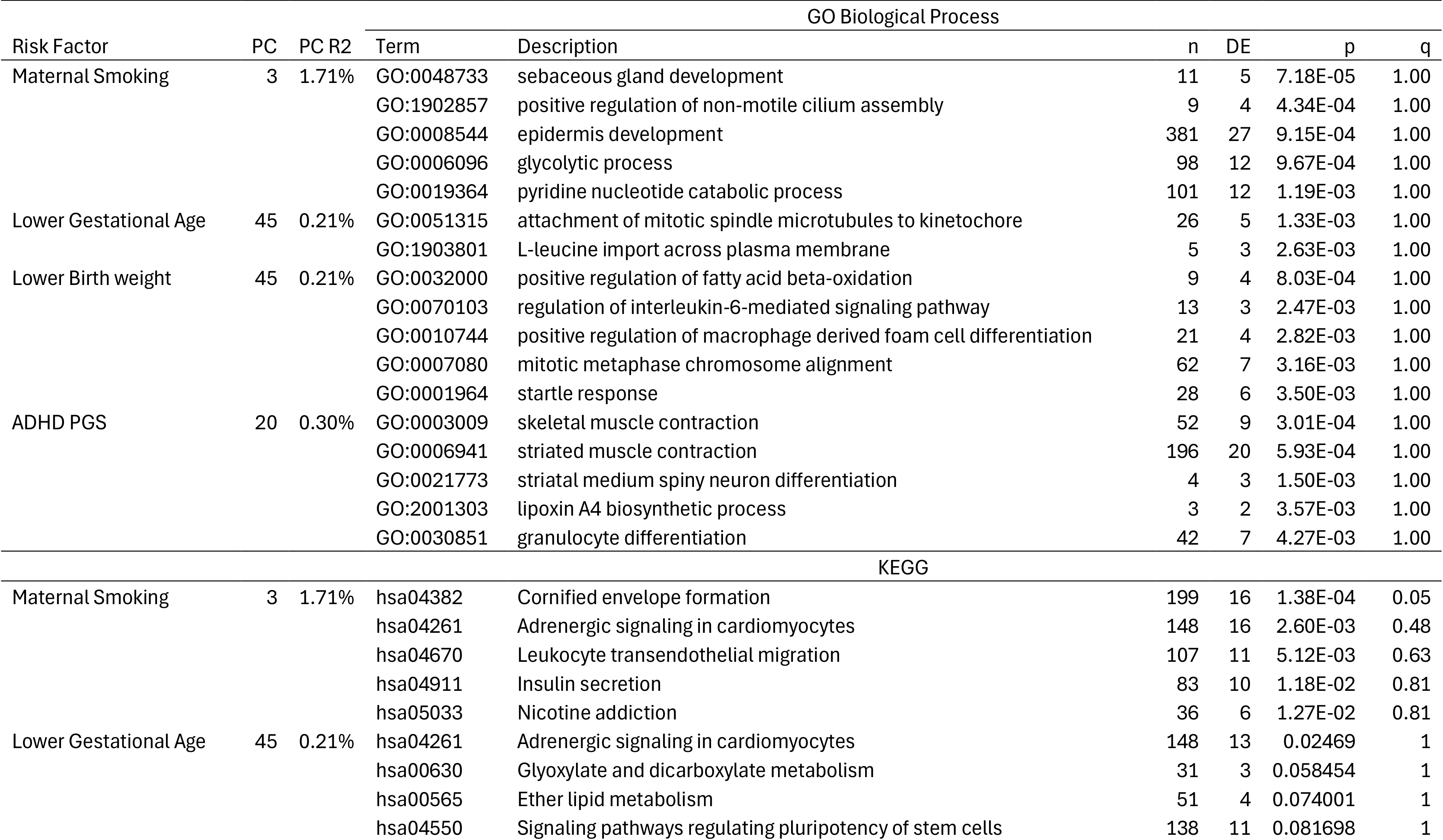
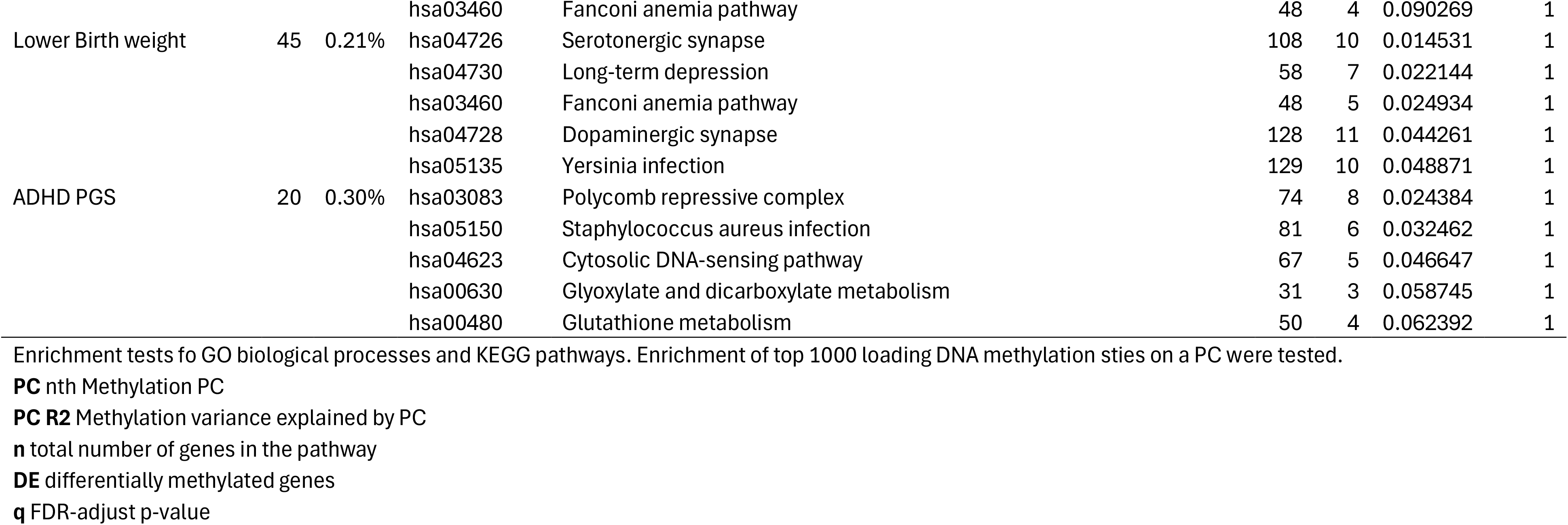
Mediation Pathways.

Birth weight effects on ADHD were globally mediated by DNAm (β_indirect_=0.025, [0.001, 0.050], p=0.045) with a proportion mediated of 55.6%. The top PC contributing to this global effect was PC45 (β_indirect_=0.008, [0.003, 0.013], p=1.3*10^-3^, q=0.13) with a proportion mediated of 17.8%, but it did not survive multiple testing correction. Top nominally significant KEGG pathways included serotonergic (p=0.01, q=1.00) and dopaminergic synapses (p=0.04, q=1.00).

The PCMA results also suggest that genetic predisposition for ADHD is mediated via DNAm with a proportion mediated of 35.2% (β_indirect_=0.062, [0.035, 0.090], p=8.4*10^-6^). Two PCs showed nominally significant associations: PC20 (β_indirect_=0.010, [0.002, 0.018], p=0.011, q=0.54) and PC45 (β_indirect_=0.009, [0.002, 0.016], p=0.011, q=0.54). As PC loadings are determined conditional on the exposure, the PC45 for ADHD-PGS moderately differs from the birth weight PC (r=0.70). Enrichments tests for PC20 primarily highlight motor neuron development with muscle contraction, as well as spiny neuron and granulocyte differentiation, among the top pathways (Table 5).

Gestational age showed nominally significant mediation via multiple DNAm PCs. However, these had inconsistent direction of effects, which when summed into a global mediation effect result in an overall null global mediation estimate. The top PC45 showed the expected mediation effect direction with a proportion mediated of 24.8%, which was FDR significant (β_indirect_=0.011, [0.005, 0.016], p=1.4*10^-4^, q=0.01). This PC45 correlated 0.86 with the gestational age PC45 and 0.65 with the ADHD-PGS PC45.

Pre-pregnancy BMI did not show evidence for mediation via DNAm in any of the cohorts. Individual PCs also did not show nominally significant mediation effects.

In sensitivity analyses, we repeated PCMA mediation analyses for the environmental exposures, but now adjusting all mediation pathways for the ADHD-PGS. Results were very similar to the main analyses (Figure 3, Table S3). Notably, mediation via PC45 remained nearly unchanged for gestational age and birth weight (Table S4).

**Figure 3:**
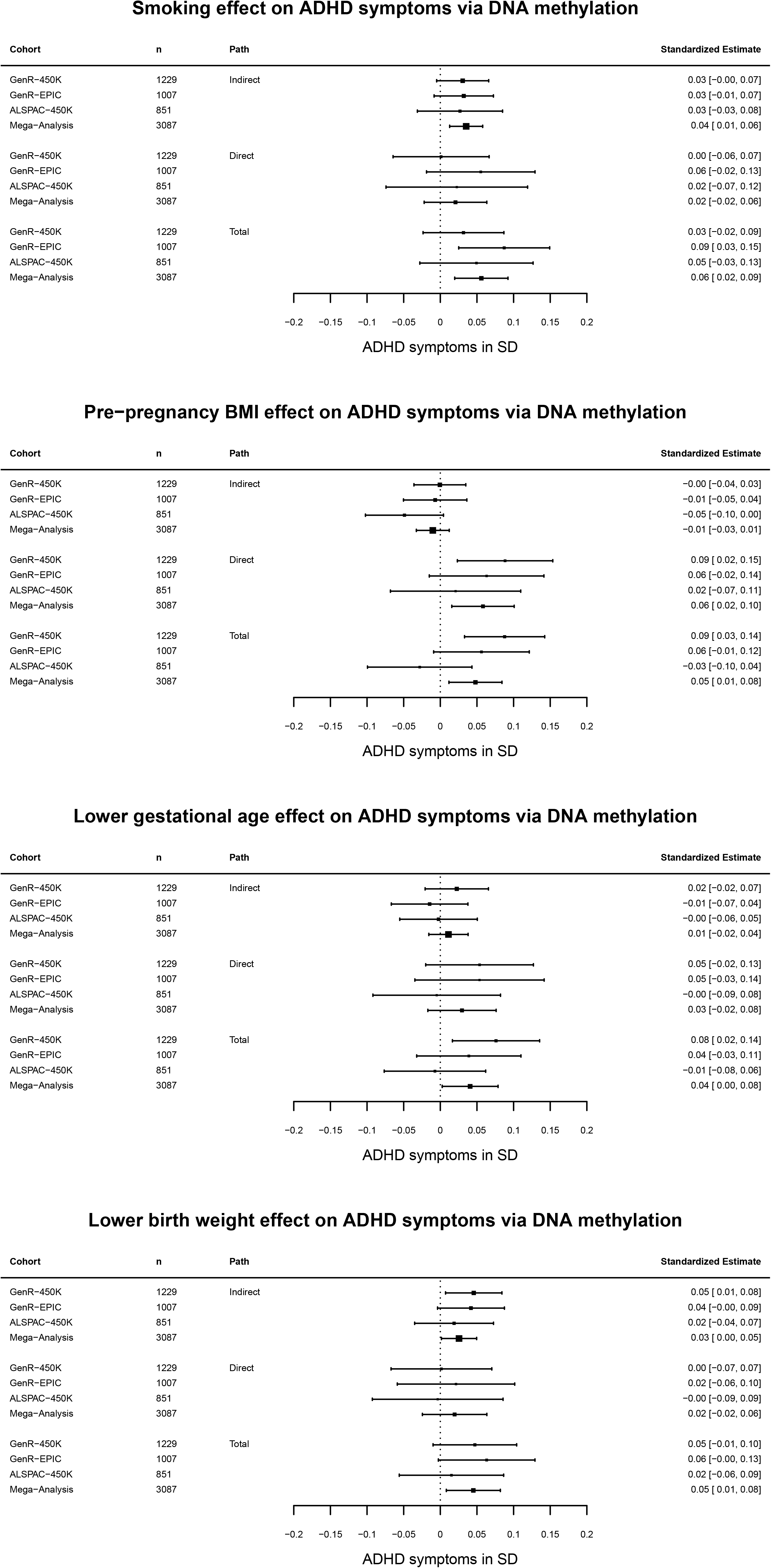
Global mediation estimates of perinatal risk factors on ADHD symptoms adjusted for ADHD-PGS. Global mediation estimates are based on PCMA analyses. They represent the standardized effect of a perinatal risk factor via 100 DNAm PCs, which themselves are based on 18,387 DNAm sites nominally associated with ADHD symptoms. **“**Indirect” path refers to the mediated effect (a*b), “Direct” to the non-mediated pathway (c’) and “Total” to the effect of both pathways (a*b+c’). Estimates are provided across the whole study sample (Mega-analysis), as well as stratified by (sub-)cohort. Analyses were adjusted for child age, sex, maternal education, maternal age, national origin, cohort and ADHD-PGS. DNAm was residualized for sample plate, array, and cohort.

For further context, we tested enrichment for all 18,387 DNAm ADHD-associated DNAm sites, as opposed to top 1000 loading for an individual PC. These analyses indicate neuronal developmental pathways such as neuron projection and axon guidance, positive regulation of nervous system development, hindbrain development, but none reached FDR significance (Table S5).

Finally, we also tested the mediation effects of single DNAm sites using traditional mediation models. Using a DACT test for mediation, p-value distributions showed no major signs for deflation or inflation (Figure 4). However, no individual DNAm site reached genome-wide significance. We also performed a look-up of GFI1 DNAm sites previously reported to mediate maternal smoking effects (Miyake et al., 2021). While DNAm levels in cg12876356 and cg18146737 were lower for children exposed to maternal prenatal smoking, DNAm levels were not associated with ADHD symptoms (Table S6). Full summary statistics and PCMA output can be found at doi.org/10.5281/zenodo.21531340.

**Figure 4:**
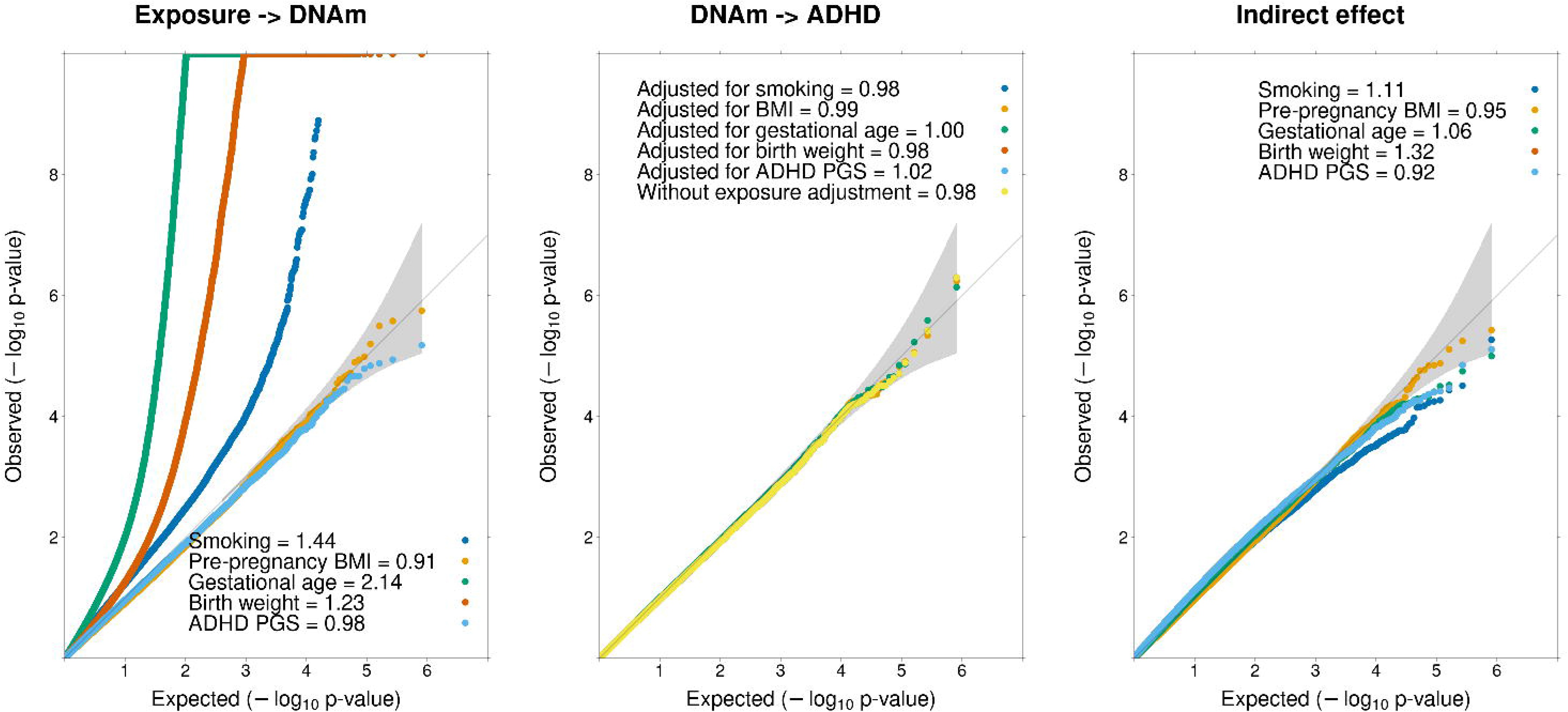
QQ-plots. The figure displays the observed vs expected p-value distribution of single DNAm site analyses conducted genome-wide. Grey area indicates 95% confidence interval. P-values were derived from a traditional mediation analysis using SEM (MLR) with a DACT test for indirect effects. P-values below 1×10^-10^ were winsorized. Analyses were adjusted for child age, sex, maternal education, maternal age, national origin, and cohort. DNAm was residualized for sample plate, array, and cohort. The left panel displays the p-value distribution for the a pathway (perinatal risk factor to DNAm). The middle panel shows the b pathway (DNAm to ADHD symptoms). “Without exposure adjustment” refers to an EWAS of ADHD symptoms without mediation effects performed for screening purposes. The right panel represents the mediated effect (a*b). Additionally, inflation lambda values are presented for each analysis.

**Figure 5:**
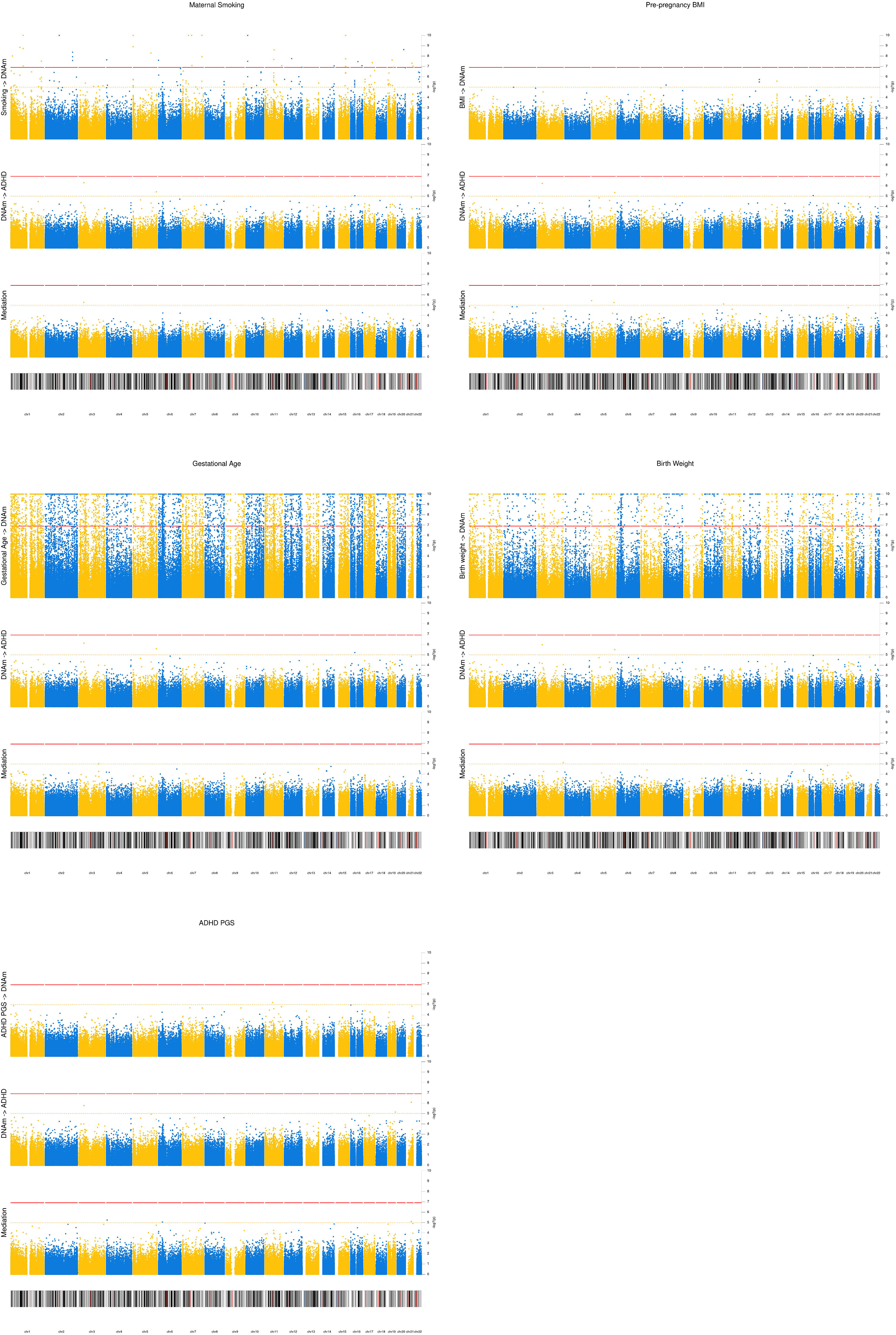
Manhattan plot. Manhattan plots showing -log10 p-values (y-axis) for the a pathway from perinatal risk factor to DNAm (top), the b path from DNAm to ADHD symptoms (middle) and the mediated path (a*b, bottom) per site (x-axis) per perinatal risk factor. P-values were derived from a traditional mediation analysis using SEM (MLR) with a DACT test for indirect effects. P-values below 1×10^-10^ were winsorized. Analyses were adjusted for child age, sex, mat. education, mat. age, national origin, and cohort. DNAm was residualized for sample plate, array, and cohort. Red line represents Bonferroni threshold (1.2*10^-7^) and yellow line suggestive threshold (1*10^-5^). Bottom ideogram displays cytobands.

## Discussion

In this study we explored whether the effects of perinatal and genetic risk factors on child ADHD symptoms is mediated by DNAm at birth. Across two large population-based birth cohorts we replicated previously reported associations of maternal smoking, higher pre-pregnancy BMI, lower gestational age, lower birth weight, and higher ADHD-PGS with greater ADHD symptoms assessed longitudinally in childhood. Using a high-dimensional mediation approach we found DNAm statistically mediating the relationship of maternal smoking and low birthweight with ADHD symptoms, with DNAm explaining about half of the total effect. Furthermore, results suggest a role of DNAm in mediating genetic predisposition for ADHD, mediating about a third of the effects. Evidence for mediation of pre-pregnancy BMI and gestational age effects on ADHD was either weak or inconsistent.

Among the environmental factors, prenatal maternal smoking had the strongest association with ADHD and most evidence for mediation via DNAm, with a proportion mediated of 62%. Several causal mechanisms have been proposed explaining the link between maternal smoking and ADHD development, including direct effects on the fetal nervous system, e.g. via nicotinic acetylcholine receptors, or more indirectly by contributing to hypoxia and ischemia (Ekblad et al., 2015; Tsegay et al., 2026). However, smoking may also represent familial confounding, e.g. by inherited genetic variants pleiotropically affecting both smoking and ADHD. Previous meta-analyses suggest that associations persist after adjustment for maternal age, socioeconomic status, alcohol consumption and ADHD history (Zakerinasab et al., 2026), supporting (but not confirming) a causal effect. Evidence from mendelian-randomization (MR) (Sorjonen & Melin, 2024; Xie & Mao, 2024), sibling (Obel et al., 2016) and egg donation studies (Thapar et al., 2009) trend towards non-causal interpretations. To examine the potential of genetic confounding, we performed secondary analyses adjusting mediation models for the ADHD-PGS. This had only a minor impact, reducing both the indirect and total effect by less than 12%. Our results therefore do not support the hypothesis that maternal smoking effects are driven by genetic susceptibility to ADHD, but as the PGS does not capture all possible genetic effects, residual genetic confounding remains a possibility.

The large mediation estimate suggests that cord blood DNAm either itself causally mediates prenatal smoking effects on ADHD, is a good proxy for other causal mechanisms, e.g. brain DNAm, or confounded by non-epigenetic factors. The lack of significant individual DNAm-PCs makes biological interpretation difficult and may hint at involvement of several mechanisms with low effect, one of which may be related to skin development according to nominal enrichment tests. A previous candidate study suggested mediation of maternal smoking effects on ADHD through *GFI1* DNAm (Miyake et al., 2021). We attempted to replicate the effect of two *GFI1* sites also available in this study, but found them to be not associated with ADHD symptoms.

In addition to prenatal smoking, we also estimated a substantial DNAm mediated effect for birth weight, with 47% mediated. Evidence for a causal effect of birth weight on ADHD is substantially stronger than that of maternal smoking. Twin studies show that the lighter twin is at higher risk of developing ADHD (Lim et al., 2018) and MR analyses also support causal interpretations (Orri et al., 2021). Low birthweight may represent fetal hypoxia and ischemia (Kinzler & Vintzileos, 2008; Smith et al., 2016) or inadequate nutrient intake (Prada & Tsang, 1998), which could negatively affect brain development. While not surviving multiple testing, the 45^th^ methylation PC is notable for being the top mediating individual PC for birth weight, gestational age, and the ADHD-PGS. While PC loadings are conditional on exposure, the different PC45 variants nevertheless substantially correlated. This raises the question, whether this DNAm component represents a common mediating mechanism for multiple environmental and genetic risk factors, e.g. modulation of serotonergic and dopaminergic pathways as indicated by nominal enrichment tests. Alternatively, the results for birth weight and gestational age may be confounded by genetic effects, however, adjusting environmental mediation models for the ADHD-PGS had negligeable impact.

While gestational age was the only risk factor with a significant individual PC (PC45), we did not find evidence for global mediation. Global estimates are the summation of all individual PC mediation estimates, which differed in direction resulting in a net null mediation. This may reflect statistical imprecision and instable individual estimates, but compensatory mechanisms are plausible, too. For pre-pregnancy BMI we did not see evidence that prenatal risk effects on ADHD are mediated via DNAm on either global or individual PC levels. For all risk factors, we were not able to identify individual DNAm sites mediating risk effects.

Finally, we also examined the role of DNAm in mediating genetic effects and observed the largest absolute total and mediated effect sizes for the ADHD-PGS. The ADHD-PGS had approximately three times higher association with ADHD symptoms than the environmental risk factors and the largest indirect effect. However, the relative proportion mediated was somewhat lower with 35%, perhaps as consequence of more brain-specific pathways and less systemic effects reflected in blood DNAm compared to environmental exposures. Enrichment analyses suggested the involvement of motor development, as well as differentiation of striatal spiny neurons and granulocytes. We have previously found that both ADHD genetic predisposition and cord blood DNAm are related to neurodevelopment (Serdarevic et al., 2020, 2023), and in turn, that deficits in neuromotor development are associated with the development of ADHD symptoms (Mostofsky et al., 2006; Valera et al., 2010). Genetic risk factors may therefore affect ADHD by epigenetically-mediated deficits in neuromotor development, but as enrichments tests were not significant after multiple testing, this hypothesis requires further replication.

Strengths of this study include the use of formal mediation tests and use of high-dimensional mediation models, which have not been applied in the context of DNAm and ADHD before. Another strength was the mega-analysis design with two population-based cohorts, totaling over 3000 children. The large sample size allows for more confident estimation of DNAm PCs and sufficient power for mediation testing of modest perinatal risk effects. Cohort and array stratified analyses suggest largely consistent estimation of mediated effects for maternal smoking, birth weight and ADHD-PGS and support the generalizability of the identified global effects despite different populations, differences in DNAm and ADHD ascertainment, and other study differences. Finally, the integration of genetic risk factors allowed for better control of genetic confounding.

Power was much more limited to detect mediation effects for individual DNAm PCs or DNAm sites, despite the use of specialized well-powered tests. The higher statistical precision of the global mediation effects highlights the advantages of aggregating multiple DNAm sites in the study of epigenetic mediation. Furthermore, pathway enrichment did not survive multiple testing correction, thus we can only carefully speculate about the exact biological mechanisms underlying the global mediation effects. Interpretation of biological mechanisms is further complicated by the observational design. While we carefully controlled each mediation model path for potential confounders, we cannot exclude the possibility of residual confounding, such as genetic effects not captured by the ADHD-PGS. The perinatal exposures could also be proxies for other risk factors and cord blood DNAm may not causally mediate effects but rather be a marker of brain DNAm or of non-epigenetic mediating factors. Imbalances in measurement error in exposure vs DNAm ascertainment may also bias mediation estimates (Richmond et al., 2016). Finally, in the case of birth weight and gestational age, reverse causality between the risk factors and DNAm could also be at play.

In conclusion, this study estimates that approximately half of the risk association of maternal smoking and lower birth weight with ADHD symptoms could be explained by DNAm at birth, even after adjusting for genetic predisposition to ADHD. Furthermore, DNAm mediated approximately one third of genetic risk effects on ADHD symptoms. In contrast, no consistent evidence for mediation was found for pre-pregnancy BMI and gestational age, suggesting that the tested DNAm sites neither causally mediate these risk effects themselves, nor correlate with other potential causal factors. We recommend further study into the role of DNAm in mediating maternal smoking, birth weight and genetic factors, and identifying the specific causal biological mechanisms. We also encourage the application of high-dimensional mediation models for other (psychiatric) outcomes and exposures and use of formal mediation testing in epigenetic epidemiology.

## Data availability

PCMA output and summary statistics can be found at doi.org/10.5281/zenodo.21531340. Analysis code can be found at github.com/aneumann-science/perinatal_mediation.

The GenR datasets generated and analyzed during the current study are not publicly available, but are available on reasonable request to the head of the Generation R Study.

The informed consent obtained from ALSPAC (Avon Longitudinal Study of Parents and Children) participants does not allow the data to be made available through any third party maintained public repository. Supporting data are available from ALSPAC on request under the approved proposal number, B3361. Full instructions for applying for data access can be found here: http://www.bristol.ac.uk/alspac/researchers/access/. The ALSPAC study website contains details of all available data (http://www.bristol.ac.uk/alspac/researchers/our-data/).

## Acknowledgements

This work was supported by the European Research Council (Tempo; grant agreement No 101039672; AN, CAMC) and European Union’s Horizon Europe Research and Innovation Programme (FAMILY, grant agreement No 101057529, AN, CAMC); HappyMums, grant agreement No 101057390, CAMC).

Views and opinions expressed are however those of the author(s) only and do not necessarily reflect those of the European Union, or the European Research Executive Agency (REA), the SERI or the UKRI. Neither the European Union nor the granting authorities can be held responsible for them.

## AI statement

The Authors declare that they have not used AI in any capacity that would require disclosure in accordance with Wiley’s AI Guidelines, nor in any capacity that would reasonably require discloser for editors, reviewers and readers to properly evaluate their research or manuscript. The authors confirms that AI has not been used for purposes including but NOT limited to drafting and editing, to generate substantial text or restructure arguments, nor in the research methodology. The authors take full responsibility for the accuracy of this statement.

## Abbreviations

ADHD: Attention-deficit/hyperactivity disorder
ALSPAC: Avon Longitudinal Study of Parents and Children
BMI: Body mass index
DACT: Divide-Aggregate Composite-null Test
DNAm: DNA methylation
EWAS: Epigenome-wide association study
GenR: Generation R Study
PCMA: Principal component mediation analysis
PGS: polygenic score

