## Supplemental information for "Perinatal risk factors, DNA methylation and the development of ADHD symptoms: a high-dimensional mediation analysis"

### Supporting Methods

#### ALSPAC ethical approval

Ethical approval for the study was obtained from the ALSPAC Ethics and Law Committee and the Local Research Ethics Committees. Informed consent for the use of all data collected was obtained from participants following the recommendations of the ALSPAC Ethics and Law Committee at the time. Participants can contact the study team at any time to retrospectively withdraw consent for their data to be used. Study participation is voluntary and during all data collection sweeps, information was provided on the intended use of data.

Biological samples are collected in accordance with the Human Tissue Act (2004). Specific Research Ethics Committee approval is sought for the consenting process at each collection sweep. Written consent, including permission for future use, is obtained from adult participants or from the parents of children as appropriate. Ethical approval for future use is covered by ALSPAC's Research Tissue Bank approval.

All historical consents to hold biological samples have been reviewed as part of the Tissue Bank approval process. Participants can contact the study team at any time to retrospectively withdraw consent for use of their samples

#### Maternal Smoking

Maternal smoking behavior during pregnancy was assessed with questionnaires (Duijts et al., 2008; Richmond et al., 2014). We categorized maternal smoking as any versus no smoking during pregnancy, see supporting information. While sustained smoking has larger effects on DNAm (Joubert et al., 2016), associations with ADHD did not differ between sustained smoking or stopping smoking in early pregnancy in this study (Table S1). As merging smoking categories increased precision of total effects and simplified statistical modeling, we present the results for any smoking during pregnancy.

#### Gestational Age

Gestational age was based on ultrasound in GenR using biparietal diameter (n=1472), crown–rump length (n=818), abdominal transverse diameter (n=41) or derived from medical records (n=2) (Verburg et al., 2008). In ALSPAC, gestational age was extracted from birth records, which were most often based on last menstrual period. Birth weight was also obtained from birth records in both cohorts.

#### ADHD-PGS

We used an existing PGS from previous work (Schuurmans et al., 2025). Briefly, genotyping was performed with Illumina HumanHap 610, 660 Quad and GSA-MD v2.0

chips in GenR and imputed to 1000G Phase IIIv5 (Ghatan et al., 2025; Medina-Gomez et al., 2015; The 1000 Genomes Project Consortium, 2015). In ALSPAC, Illumina 550 chips were used for genotyping and imputed to 1000G Phase Iv3. For PGS calculations we only used autosomal SNPs with at least 1% minor allele frequency and 80% imputation quality. The PGS was based on a ADHD case-control GWAS (Demontis et al., 2023) and computed using a clumping and thresholding approach with PRSice-2 (Choi & O'Reilly, 2019). A p-value threshold of 0.01 was chosen, at it had previously the most consistent performance between cohorts (Schuurmans et al., 2025). The PGS was z-scored and residualized for twenty genomic principal components to account for population stratification.

#### DNA methylation

DNAm was measured in cord blood with either the Illumina HumanMethylation450 BeadChip array in GenR (GenR-450K) and ALSPAC (ALSPAC-450K) or MethylationEPIC v1.0 BeadChip in GenR only (GenR-EPIC). The Zymo EZ-96 DNAm kit was used for bisulfite conversion. In ALSPAC the meffil package (Min et al., 2018) was used for DNAm processing with quality control including mismatched genotypes or sex, relatedness, low concordance of DNAm levels with other time points, extreme dye bias, and poor probe detection. In GenR the CPACOR workflow was applied (Lehne et al., 2015). Quality control included checks for bisulfite conversion, hybridization or extension, sex mismatches and sample call rate of 95% (450K) or 96% (EPIC). DNAm values were functionally normalized in meffil using ten control probe principal components and slide as a random effect. GenR-450K and ALSPAC-450K were previously normalized as combined dataset (Mulder et al., 2021). Further harmonization with GenR-EPIC is described below.

We restricted the DNAm matrix to autosomal sites present on both arrays. We excluded cross-reactive probes using the maxprobes package(Chen, 2018/2024). Methylation values were converted to M-values. We merged the GenR-450K, GenR-EPIC and ALSPAC-450K datasets and residualized M-values on random effects of sample plate, array and cohort to reduce technical and population-specific effect, as PCMA does not support covariate inclusion in the analysis model.

### Additional acknowledgements

#### Generation R

The Generation R Study is conducted by Erasmus MC, University Medical Center Rotterdam in close collaboration with the School of Law and Faculty of Social Sciences of the Erasmus University Rotterdam, the Municipal Health Service Rotterdam area, Rotterdam, the Rotterdam Homecare Foundation, Rotterdam and the Stichting Trombosedienst & Artsenlaboratorium Rijnmond (STAR-MDC), Rotterdam. We gratefully acknowledge the contribution of children and parents, general practitioners, hospitals, midwives and pharmacies in Rotterdam. The generation and management of the Illumina 450K methylation array data (EWAS data) for the Generation R Study was executed by the Human Genotyping Facility of the Genetic Laboratory of the Department of Internal Medicine, Erasmus MC, the Netherlands. We thank Mr. Michael Verbiest, Ms. Mila Jhamai, Ms. Sarah Higgins, Mr. Marijn Verkerk and Dr. Lisette Stolk for their help in creating the EWAS database. We thank Dr. A.Teumer for his work on the quality control and normalization scripts.

The general design of the Generation R Study is made possible by financial support from the Erasmus MC, Erasmus University Rotterdam, the Netherlands Organization for Health Research and Development and the Ministry of Health, Welfare and Sport. The EWAS data were funded by a grant from the Netherlands Genomics Initiative (NGI)/Netherlands Organisation for Scientific Research (NWO) Netherlands Consortium for Healthy Aging (NCHA; project nr. 050-060-810), by funds from the Genetic Laboratory of the Department of Internal Medicine, Erasmus MC, and by a grant from the National Institute of Child and Human Development (R01HD068437).
JF received a Helmholtz International Fellow Award HIFA-0174 -RA-30/19. This project has received funding from the European Union's Horizon Europe Research and Innovation Programme under grant agreement nº 101137146 (STAGE) Views and opinions expressed are however those of the author(s) only and do not necessarily reflect those of the European Union. Neither the European Union nor the granting authority can be held responsible for them.

The generation and management of GWAS genotype data for the Generation R Study was done at the Human Genomics Facility, HuGe-F, housed within the Laboratory for Population Genomics of the Department of Internal Medicine at Erasmus MC. Genetic Laboratory of the Department of Internal Medicine, Erasmus MC, The Netherlands. We thank Zahra Alawi, Marijn Verkerk, Dr. Katerina Trajanoska, Costanza Vallerga, Samuel Gathan, Dr. Carolina Medina-Gomez, Dr. Linda Broer and Jard de Vries for their help in creating, managing and QC the GWAS database.

#### ALSPAC

We are extremely grateful to all the families who took part in this study, the midwives for their help in recruiting them, and the whole ALSPAC team, which includes data collection staff, data and administrations staff, technical managers and the technical staff with the Bristol Bioresource Laboratory, based within the University of Bristol.

The UK Medical Research Council and Wellcome (Grant ref: MR/Z505924/1) and the University of Bristol provide core support for ALSPAC. This publication is the work of the authors and Alexander Neumann will serve as guarantors for the contents of this paper.

A comprehensive list of grants funding is available on the ALSPAC website

(http://www.bristol.ac.uk/alspac/external/documents/grant-acknowledgements.pdf); This research was specifically funded by BBSRC (BBI025751/1 and BB/I025263/1), IEU (MC_UU_12013/1 & MC_UU_12013/2 & MC_UU_12013/8), National Institute of Child and Human Development grant (R01HD068437), and NIH (5RO1AI121226-02 and 5R01MH073842-04).

Genomewide genotyping data was generated by Sample Logistics and Genotyping Facilities at Wellcome Sanger Institute and LabCorp (Laboratory Corporation of America) using support from 23andMe.
